# Genetically determined serum testosterone level and Covid-19 illness level: A mendelian randomization study

**DOI:** 10.1101/2021.10.10.21264779

**Authors:** Juan Xiong, Xuejun Kong, Ziyong Ma, Yifei Qu, Xu Yang, Ruicheng Miao, Jie Huang

**Affiliations:** School of Public Health, Health Science Center, Shenzhen University, Shenzhen, Guangdong, China; Athinoula A. Martinos Center for Biomedical Imaging, Massachusetts General Hospital, Harvard Medical School, Charlestown, MA, USA; Jinan Foreign Language School, Jinan, Shandong, China; Shandong experimental high school, Jinan, Shandong, China; Shenzhen Aone Medical Laboratory, Shenzhen, China; Department of Global Health, Peking University School of Public Health, Beijing, China; Institute for Global Health and Development, Peking University, Peking, China

**Author notes:** **Corresponding author:** Jie Huang, M.D, PhD, Department of Global Health, Peking University School of Public Health, 38 Xueyuan Rd, Haidian District, Beijing, China.

**Keywords:** testosterone level, Covid-19, Mendelian Randomization

## Abstract

**Background:** It is hypothesized that different levels of hormones especially serum testosterone level could explain the sex differences between men and women on the susceptibility and case fatality rate of COVID-19. However, traditional observational studies that support this hypothesis could not effectively establish the causal effects.

**Objective:** Utilizing recently published genome-wide associations studies (GWAS) on serum Testosterone level and on COVID-19 related phenotypes, we sought to assess the causality through Mendelian Randomization (MR) analyses. We further applied a suite of statistical genomics methods to further explore the biological mechanisms.

**Results:** We found that testosterone level is significantly associated with Covid-19 critical illness. All six MR methods yielded significant associations. There is no significant association between Testosterone and COVID-19 respiratory failure or COVID-19 susceptibility.

**Conclusion:** Based on the GWAS currently available, we provide support for a causal role of Testosterone on COVID-19 critical illness.Nevertheless, we recognize that the COVID-19 susceptibility GWAS effort is still ongoing and there is no such strong locus as CCR5 for HIV discovered for COVID-19.

## Introduction

It has been observed that men with COVID-19 are more at risk for worse outcomes and death, independent of age.^1^ Furthermore, the case fatality rate in men is twice of that in women. One theory is that different levels of hormones especially serum testosterone could explain. However, it is apparent that sex difference is driven by and associated with many more other (risk) factors. Height is an obvious one: although height is also reported to be associated with Covid-19 susceptibility, the association is far too weak to explain the 2:1 ratio of case fatality between men and women.

Within males, it is reported that low testosterone levels in the blood are linked to more severe progression of Covid-19.^2^ Still, these types of studies could not conclude that this association is causal. While it is plausible that testosterone plays a protecting role, it was also reported that genetically predicted endogenous testosterone is detrimental for thromboembolism, heart failure, and myocardial infarction, especially in men^3^. Furthermore, observational studies could not tease apart reverse causation (the SARS-COV-2 virus induces an acute reduction in testosterone levels) and confounding effects (the low levels of testosterone could also be a marker of some other causal factors).

Biologically, a high level of testosterone results in the upregulation of angiotensin-converting enzyme type 2 (ACE2) and transmembrane protease serine 2 (TMPRSS2) receptors.^4^ Testosterone could affect both susceptibility to infection and disease severity through its effect on these two.^5,6^ Low levels of testosterone deregulates lung-protective pathways.^7,8^ Also, age-associated decreases in sex steroid hormones (estrogen and testosterone) may mediate proinflammatory increases in older adults that increases their risk of COVID-19 adverse outcomes.^4^

Therefore, it is imperative to assess the causal role of testosterone on Covid-19 progression. This will provide insights to ongoing clinical trials that investigates hormonal therapies that block or lower testosterone or increase estrogen as a treatment for Covid-19 patients. In this study, we apply mendelian randomization. We used 35 biomarkers, including estradiol IGF-1. We first used summary statistics data, and then we used individual level data. We run the GWAS for males and females separately, and we adjust for age, body mass index (BMI), and the other 34 biomarkers.

## Methods

### Serum testosterone level GWAS

We downloaded the Testosterone GWAS summary statistics from the Sinnott-Armstrong study, which studied and made publicly available 35 blood and urine biomarkers in the UK Biobank^9^. We merged the array data and the imputed data together. For the imputed data that does not come with rsID, we used a customized add_rsid module from pheweb to add_rsID. There are ∼9 million SNPs for the merged data.

### Covid-19 GWAS

We downloaded three COVID-19 GWAS summary statistics from GWAS Catalog: (1) COVID-19 critical illness^10^, (2) severe COVID-19 with respiratory failure^11^, (3) COVID-19 susceptibility^12^. For those without rsID, we used the add_rsID module mentioned above to add rsID.

### Mendelian Randomization analysis

We used 113 independent lead SNPs associated with Testosterone level (*P* <5E-09) as instrumental variables.^9^ Among them, 1, 12, 100 are protein-truncating, protein-altering, non-coding variants respectively. We extracted the summary statistics of these SNPs from the Testosterone GWAS and the COVID-19 GWAS. For each pair (Testosterone vs. one of the 3 COVID-19 GWAS), we merge the summary statistics and then align the effect to the same allele. We then used the Mendelian Randomization R package^13^ to conduct a 2-sample MR, where Testosterone level is exposure while COVID-19 is outcome.

### Functional analysis

For all four GWAS, we used FUMA software^14^ to prioritize the set of genes associated with each trait. FUMA deals with the fact that GWAS results typically do not directly translate into causal variants because the majority of hits are in non-coding or intergenic regions, and the presence of linkage disequilibrium (LD) leads to effects being statistically spread out across multiple variants. It accommodates positional, expression quantitative trait loci (eQTL) and chromatin interaction mappings, and provides gene-based, pathway and tissue enrichment results. We further used a universal enrichment tool to perform enrichment analyses and generating plots^15^.

## Results

The Manhattan plot Testosterone GWAS was shown in **Figure 1**. The instrumental variants were highlighted in green color. Of note, not all top SNPs chose to be the instrumental variables have the lowest P-values. We found that testosterone level is significantly associated with Covid-19 critical illness, as defined by Pairo-Castineira. *et al*.^10^ **Figure2** shows the Mendelian Randomization results between Testosterone level and COVID-19 critical illness. All six methods yield significant associations. There is no significant association between Testosterone and COVID-19 respiratory failure or COVID-19 susceptibility. **Figure 3** shows the funnel plot of the MR analysis mentioned above.

**Figure 1.**
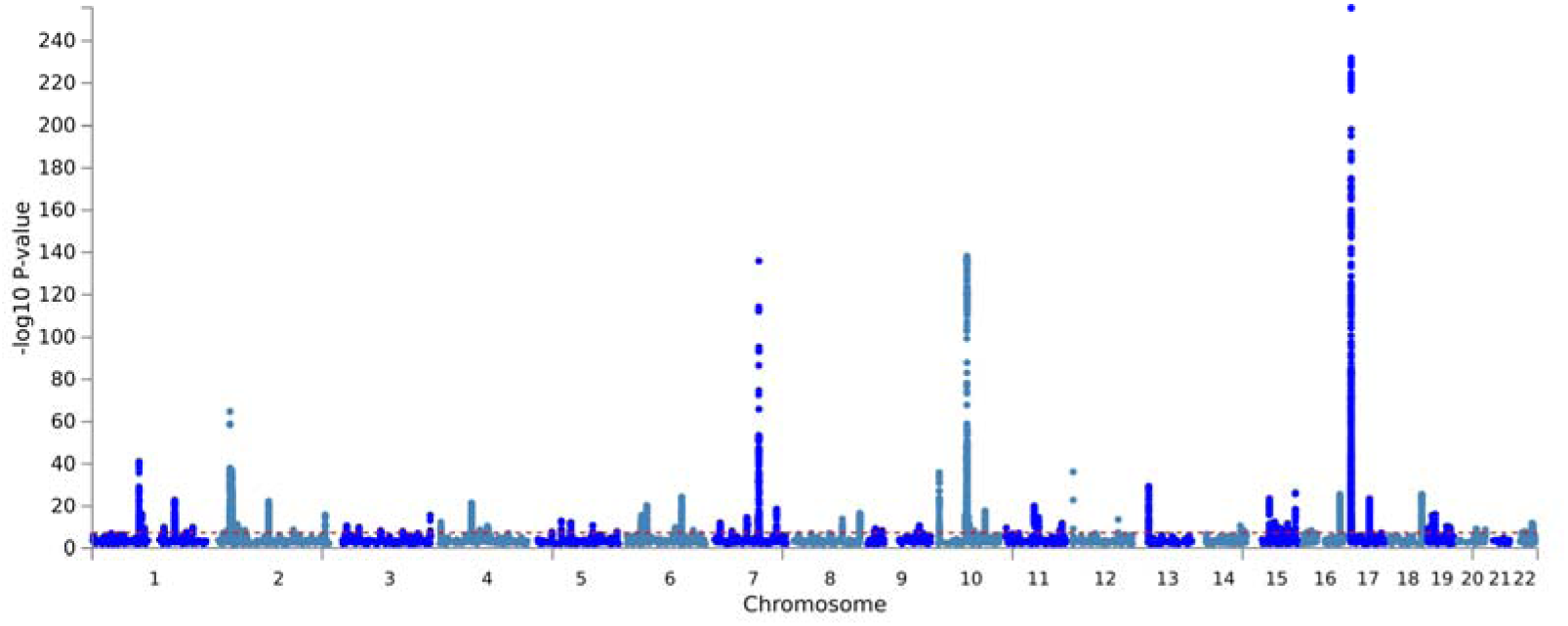
Manhattan plot of Testosterone GWAS

**Figure 2.**
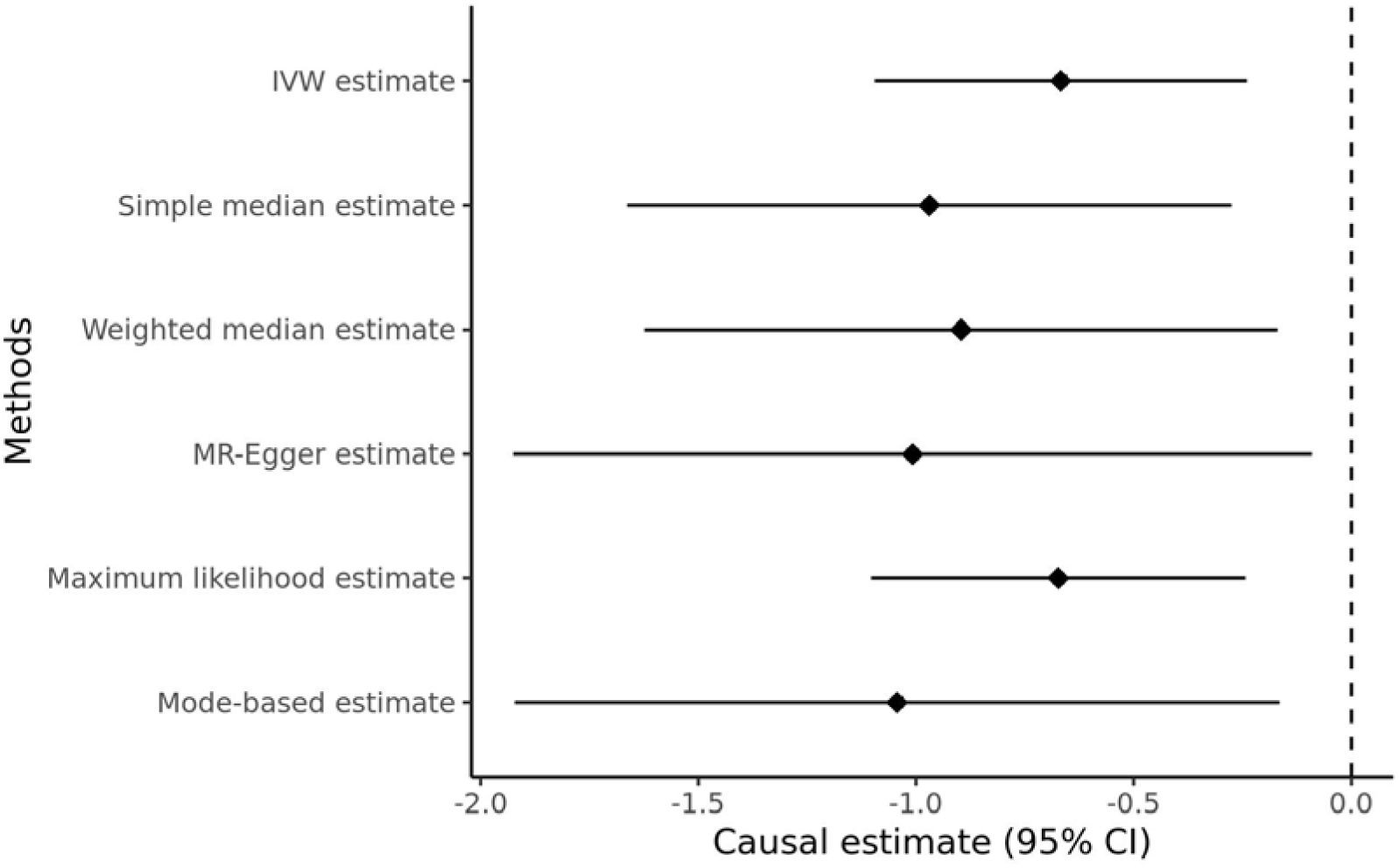
Mendelian randomization between Testosterone and Covid-19 critical illness

**Figure 3.**
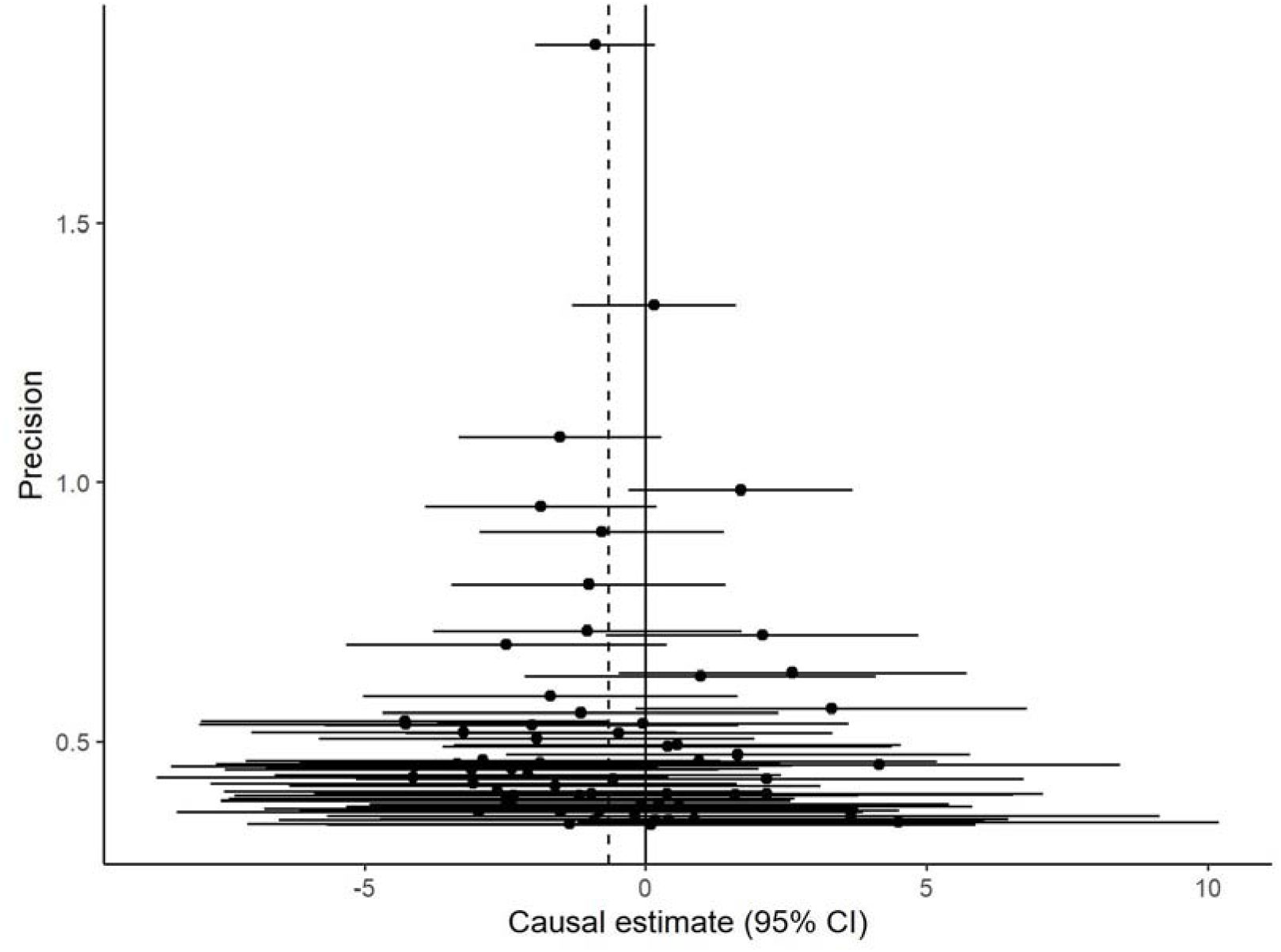
Mendelian randomization funnel plot between Testosterone and Covid-19 critical illness

**Figure 4.**
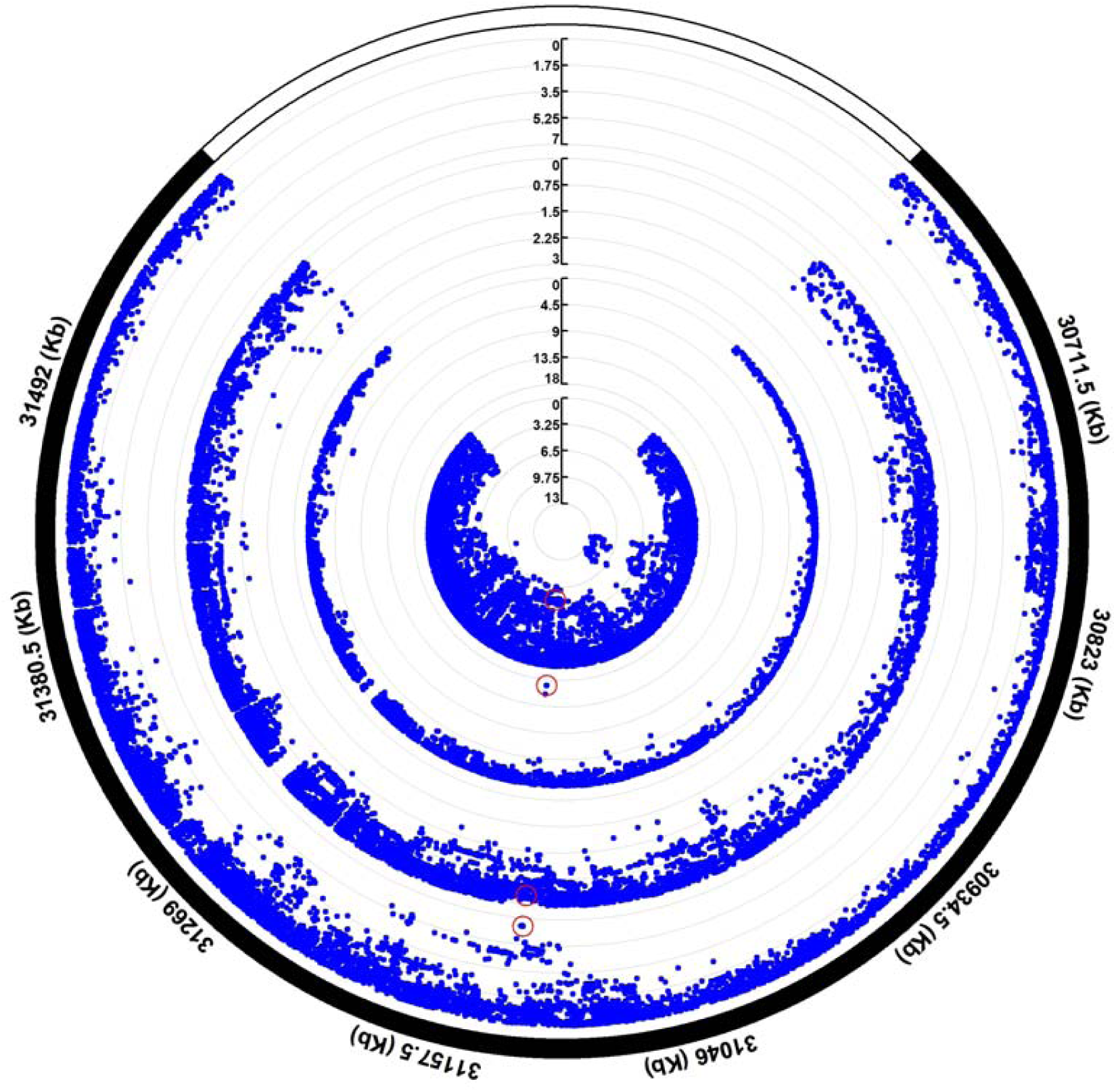
the colocalization result of *CCHCR1* region

At the individual SNP level, we identified an intronic variant on *CCHCR1* that is genome-wide significant in both Testosterone and Covid-19 ICU GWAS (**Figure 5**). The *CCHCR1* gene (Coiled-Coil alpha-Helical Rod protein 1), also simplified as HCR gene, was shown to promote steroidogenesis by interacting with the steroidogenic acute regulator protein (StAR). A recent GWAS preprint by Genetics of Mortality in Clinical Care (GenOMICC) collaborators including HGI (https://genomicc.org) reported that among a few other genes CCHCR1 is associated with COVID-19 severity.^16^ The lead SNP is rs72856706 (hg19 position chr6:31,120,729). It has a frequency of 0.08 for the non-reference allele (T), based on both TopMed and GnomAD. The association statistics are respectively: Testosterone (Beta=0.029, P=7.307E-09); COVID-ICU: (BETA=0.614, P=9.15E-18); Covid-19 susceptibility (beta=0.039, P=3.44E-07). However, it is non-significant for Covid-19 respiratory failure (Beta=0.052, P=0.58). The *CCHCR1* (HCR) gene was reported to be relevant for skin steroidogenesis.

## Discussion

We run a comprehensive set of statistical and bioinformatic analyses to assess the causal role of serum Testosterone level on COVID-19 related phenotypes. Our analyses results support a causal role. However, we realize the following limitations. First, the GWAS results on COVID-19 are inconsistent. The COVID-19 susceptibility GWAS effort is still ongoing and there is a lack of well-established loci. So, far, there is no such strong locus as CCR5 for HIV discovered for COVID-19. Although it is reasonable to hypothesize that the disease severity of COVID-19 has a strong genetic underpinning, the GWAS on these traits published so far are also inconsistent. The two GWAS used by the current study^1011^don’t have very consistent signals. Both studies only have thousands of cases, much smaller compared with many existing GWAS for other complex traits.

## Data Availability

All data produced in the present study are available upon reasonable request to the authors

